# Evaluation of health system resilience in 60 countries based on their responses to COVID-19

**DOI:** 10.1101/2022.09.25.22280337

**Authors:** Laijun Zhao, Yajun Jin, Lixin Zhou, Pingle Yang, Ying Qian, Xiaoyan Huang, Mengmeng Min

## Abstract

**Introduction:** In 2020, the COVID-19 epidemic swept the world, and many national health systems faced serious challenges. To improve future public health responses, it’s necessary to evaluate the performance of each country’s health system.

**Methods:** We developed a resilience evaluation system for national health systems based on their responses to COVID-19 using four resilience dimensions: government governance and prevention, health financing, health service provision, and health workers. We determined the weight of each index by combining the three-scale and entropy-weight methods. Then, based on data from 2020, we used the Technique for Order of Preference by Similarity to Ideal Solution (TOPSIS) method to rank the health system resilience of 60 countries, then used hierarchical clustering to classify countries into groups based on their resilience level. Finally, we analyzed the causes of differences among countries in their resilience based on the four resilience dimensions.

**Results:** Switzerland, Japan, Germany, Australia, South Korea, Canada, New Zealand, Finland, the United States, and the United Kingdom had the highest health system resilience in 2020. Eritrea, Nigeria, Libya, Tanzania, Burundi, Mozambique, Republic of the Niger, Benin, Côte d’Ivoire, and Guinea had the lowest resilience. Government governance and prevention of COVID-19 will greatly affect a country’s success in fighting future epidemics, which will depend on a government’s emergency preparedness, stringency (a measure of the number and rigor of the measures taken), and testing capability. Given the lack of vaccines or specific drug treatments during the early stages of the 2020 epidemic, social distancing and wearing masks were the main defenses against COVID-19. Cuts in health financing had direct and difficult to reverse effects on health systems. In terms of health service provision, the number of hospitals and intensive care unit beds played a key role in COVID-19 clinical care.

**Conclusion:** Resilient health systems were able to cope more effectively with the impact of COVID-19, provide stronger protection for citizens, and mitigate the impacts of COVID-19. Our evaluation based on data from 60 countries around the world showed that increasing health system resilience will improve responses to future public health emergencies.

**Key Questions:** *What is already known?:*

- According to a report by the World Health Organization, the COVID-19 epidemic placed the health systems of many countries at risk of collapse.
- At present, there is no evaluation index system to measure the resilience of each country’s health system against a pandemic, and there has been no quantitative assessment of the resilience of each country’s health system based on their responses to COVID-19.

*What are the new findings?:*

- We assessed, ranked, and quantified the health system resilience of 60 representative countries based on their responses to COVID-19 using data from 2020 on four dimensions of resilience: government governance and prevention, health financing, health service provision, and the health workforce.
- Western Europe, East Asia, North America, and Southern Oceania had better health system resilience, whereas Africa had low health system resilience, with very low health financing scores and weak health systems with structural and regional imbalances.
- Health system resilience was heavily influenced by government governance and prevention, as well as by government emergency preparedness, the stringency of their response (a measure of the number and rigor of the measures taken), and their testing capability.

*What do the new findings imply?:*

- Global health system resilience varied widely among countries, and many health systems remain weak and unprepared for another pandemic such as COVID-19. As a result, future pandemics will remain a major problem for humanity if improvements are not made by each government.
- In underdeveloped countries and regions, infectious diseases can be controlled more effectively through more efficient government governance and strict surveillance and detection measures, but achieving this depends heavily on the speed of government decision making and the level of policy formulation related to the most effective way to strengthen health systems and improve their resilience. Assistance from developed country will be essential in improving resilience.

## 1. Introduction

The COVID-19 outbreak in 2019 was a global pandemic caused by Severe Acute Respiratory Syndrome Coronavirus 2 (SARS-CoV-2).^1^ As of 14 April 2022, the World Health Organization (WHO) reported 500.2 million confirmed cases and 6.19 million COVID-19 deaths worldwide (https://covid19.who.int). In the early stages of the response to COVID-19, the performance of the health systems of different countries varied significantly.

Italy was the first European country to respond to the COVID-19 outbreak.^2^ They experienced a high mortality rate due to the aging of their population, and the large and increasing number of patients overwhelmed the health system. As of 17 March 2020, the overall mortality rate of confirmed COVID-19 patients in Italy had reached 7.2% (1625 deaths / 22512 cases), which was much higher than the 2.3% rate in China.^3, 4^ The United States was one of the most strongly affected countries.^5^ Although the United States has a robust and diverse, high-quality health system, the demand for hospital resources far exceeded the hospital capacity in many areas.^6, 7^ For example, on 10 April 2020, the demand for intensive care unit (ICU) beds in New York was 8380, but only 718 beds were available. France was one of the most strongly affected European countries due to the limited number of ICU beds and a severely under-prepared health system.^8, 9^ As of 27 October 2020, the number of COVID-19 deaths reached 35,018 (https://www.worldometers.info). The performance of these health systems in responding to COVID-19 was far below the level their health systems should have been able to attain.

In contrast, the health systems of several countries performed relatively well in responding to COVID-19. China was the first country to face the COVID-19 outbreak, but it took swift and effective measures to control the first wave of the outbreak. At the same time, it continued to donate medical supplies to countries around the world to help them combat the epidemic. Exports of critical supplies grew from 12.6% of the total global trade in these supplies in 2019 to 26.7% in 2020, ranking first in the world and 2.3 times that of the United States, which ranked second.^10^ Uruguay excelled in testing, and was one of the first countries to adopt contact tracing.^11^ It tested an average of 233.7 people per confirmed case of COVID-19, versus only 1.7 in Argentina, 1.9 in Mexico, and 3.0 in Colombia (https://ourworldindata.org/coronavirus-testing#tests-per-confirmed-case). The rapid and effective measures taken by the Vietnamese government were recognized by international organizations.^12^ Through early risk assessment, timely emergency response policies, and immediate actions taken by government departments, the domestic epidemic was controlled at a very low level, which greatly alleviated the impact on the health system.

The performance of countries in responding to COVID-19 varied greatly. What factors led to such a wide variation in the performance of health systems? In this paper, we examine the health systems of 60 representative countries around the world. We constructed an evaluation system from the perspective of resilience, and used it to evaluate the performance of health systems based on their response to COVID-19 in 2020. We then explore the causes of the differences to provide policy suggestions for countries that will improve their response to future public health emergencies.

The concept of resilience originates from the Latin word *resillo*, which is translated as “elasticity”. The earliest disciplinary definition of resilience comes from ecology.^13^ It has been gradually applied to many fields such as engineering and psychology. In all these different disciplines, the main concept of resilience reflects the ability of an individual, group, or system to absorb shocks while still maintaining its basic functions in or near their original state.^14^ Resilience is a relatively new concept in the context of health systems. Health system resilience refers to the ability of a health system to maintain its core system functions when a crisis hits, to respond effectively to the crisis, and to improve and upgrade the system based on the lessons learned during the crisis.^15^ Since the end of the 20th century, the need to build resilient health systems has gradually become one of the core issues discussed in the World Health Report.^16^ Health system resilience is an important element of system adaptability, responsiveness, and stability, so it is critical for countries to learn from their response to COVID-19 and build a more resilient system.^17, 18^

When we reviewed previous studies, we found that the concept of resilience has been applied to health system evaluations. Among this research, Ammar et al.^19^ used a case study approach to examine health system resilience in Lebanon under the impact of the Syrian refugee crisis. Ling et al.^20^ assessed health system resilience in Liberia during the Ebola crisis using semi-structured interviews and focus group discussions to support thematic analysis. Watts et al.^21^ quantitatively assessed health system resilience in 101 countries by conducting a national survey in the context of climate change. Haldane et al.^22^ used a new health systems resilience framework to qualitatively analyze the response to COVID-19 in 28 countries; they summarized four important elements of an effective national response, and made recommendations accordingly. By reviewing the literature, we found that analysis of health system resilience in response to emergencies is extremely important, and especially quantitative analysis, which can reflect the problem more intuitively. However, there is a lack of quantitative analysis on health system resilience based on the responses to COVID-19, and this has prevented us from attaining a comprehensive understanding of the response of each country. How to quantitatively analyze health system resilience in this context was therefore the focus of this paper.

In 2020, COVID-19 had a severe impact on the health system of each country. This provided a unique opportunity to assess health system resilience around the world. To quantify the health system response to COVID-19, we need to develop a quantitative evaluation system. In the present study, we rethought the WHO assessment framework for resilience and explored relevant evaluation indicators to more accurately assess the actual situation. The WHO framework consists of six components: leadership and governance, health financing, health information systems, medical technologies and products, health service provision, and the health workforce.^23^ As COVID-19 has been a public health emergency, government information is critical, so we combined the leadership and governance component with the health information systems component to produce a single composite government governance and prevention component. In addition, at the beginning of the epidemic in 2020, most countries had no specific drugs available to treat COVID-19 and no vaccines, and medical knowledge, technologies, and products for COVID-19 treatment were not mature, and did not differ greatly among countries, so we did not account for medical technologies and products.^24, 25^ Governance is not effective if there is no money to pay for health workers, medicines, and hospitals, and none of the factors can help if there is no access to them. We therefore added health financing and access to healthcare as key dimensions. Finally, because infected patients must be cared for and treated, the health workforce was clearly essential and became our final dimension. To accurately and objectively describe the real situation in each country, we defined our new assessment framework based on the abovementioned four dimensions: government governance and prevention, health financing, health service provision, and health workers.

In this paper, we describe the following aspects of our study: First, we developed a health system resilience evaluation system based on the four dimensions (government governance and prevention, health financing, health service provision, and health workforce). Second, we determined the weights of each of these indicators using a combination of weighting methods, and then calculated a composite index that let us rank the health system resilience of 60 representative countries worldwide using data from 2020, with an emphasis on the Technique for Order of Preference by Similarity to Ideal Solution (TOPSIS) method. Third, we used hierarchical clustering to group the countries based on the evaluation results and classify the health system resilience level of each country. Fourth, by calculating each country’s resilience scores for each of the four dimensions, we identified deficiencies in countries with different levels of resilience, and use these difference to support policy recommendations for strengthening the global health system’s resilience against future pandemics.

## 2. Materials and Methods

### 2.1 Study area and data sources

To provide coverage of the overall global situation in 2020, we sampled the 20 countries with the highest number of COVID-19 deaths per million citizens, the 20 countries with the lowest number of deaths, and 20 countries from different regions with intermediate death tolls and with large populations and very different health systems (Figure 1). We excluded the Middle East region because death tolls were confounded by ongoing war and by a lack of reliable statistics. The selected countries have a total population of 5.633 billion people, which represented 74.3% of the total global population in 2020; a land area of 83,470,400 km^2^, accounting for 61.4% of the total land area of all countries; and a total GDP of US$80.7 trillion in 2020, which amounted to 94.5% of the total global GDP (http://data.un.org).

**Figure 1.**
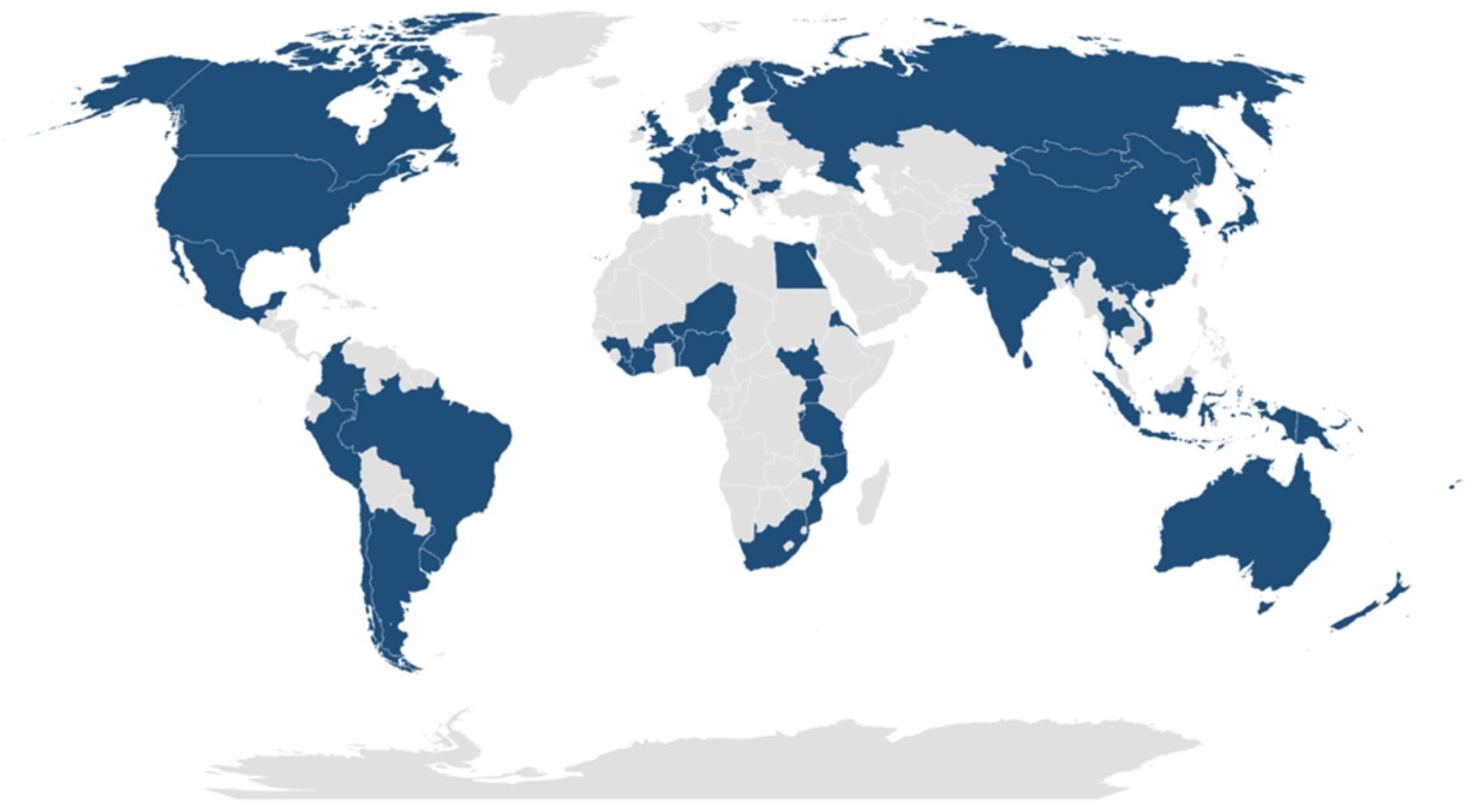
Countries included in this study.

We obtained indicator data from the latest publicly available datasets and databases produced by international organizations (WHO, the World Bank, specialized agencies of the United Nations, non-profit organizations, the Oxford COVID-19 Government Response Tracker, and the COVID-19 Regional Security Assessment Security Committee). Supplementary Table S1 describes the indicators and data sources.

### 2.2 Construction of an indicator system for assessing the resilience of health systems

By analyzing WHO’s definition of a resilient health system framework and combining it with insights from relevant studies on health system resilience, we constructed an evaluation index framework for health system resilience (Figure 2).^22, 23, 26^ We started by dividing the framework into four first-level indicators: government governance and prevention, health financing, health service provision, and health workers. We then subdivided each first-level indicator into 3 to 7 second-level indicators to facilitate more accurate qualitative and quantitative analysis.

**Figure 2.**
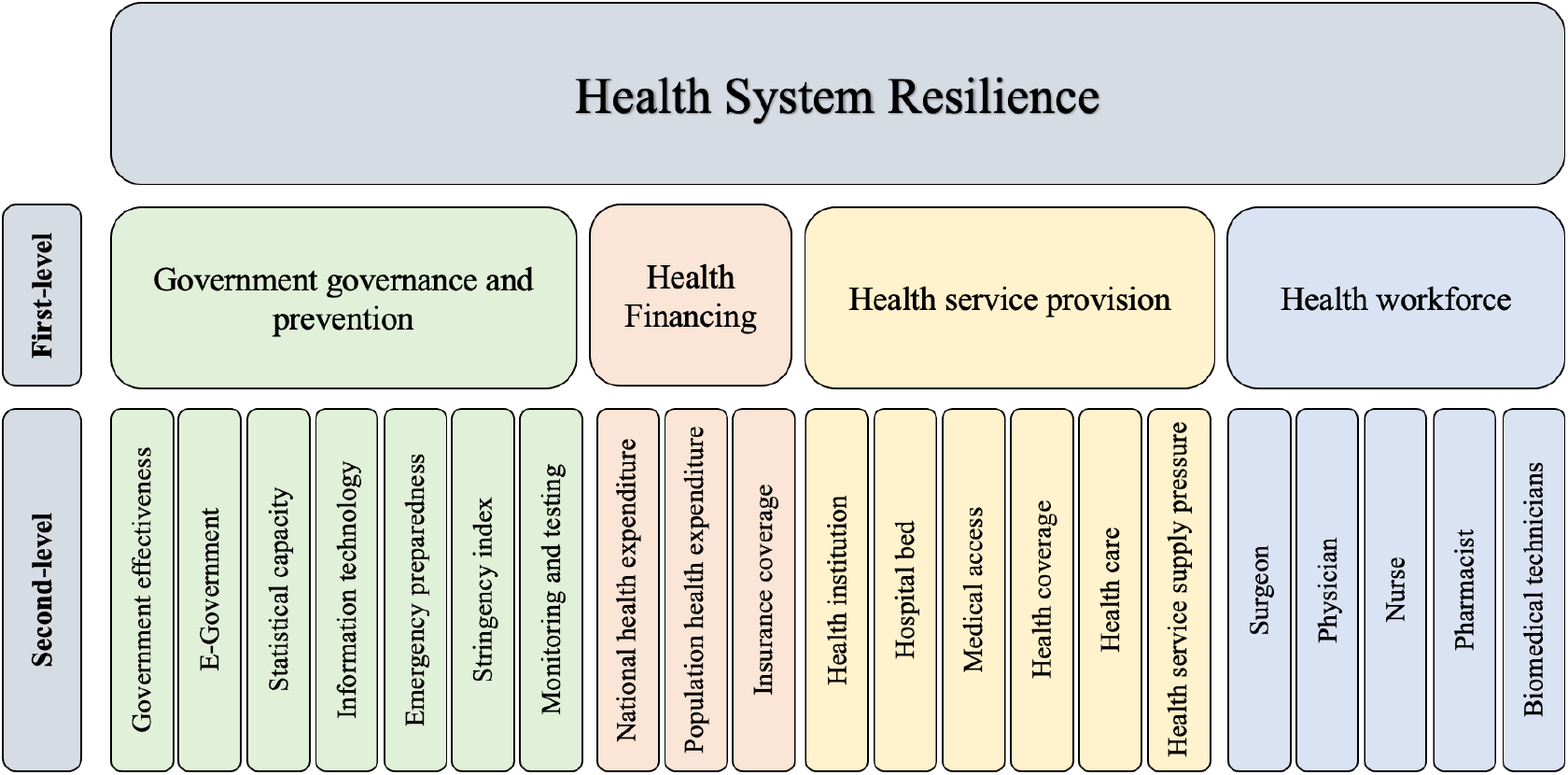
Health System Resilience framework.

#### 2.2.1 Government Governance and Prevention

Government governance and prevention is the most important factor in the health system’s response to COVID-19. The timeliness and effectiveness of the government’s response to COVID-19 and the efficiency of the formulation and implementation of relevant epidemic prevention policies will both greatly influence the development of the epidemic. The ability of the country to collect and compile data on the epidemic will affect the speed of information dissemination. The infrastructure level of the country’s information technology determines the quality of the public services the government can provide to citizens, and the sharing of information and data can guide citizens to take effective response measures.^27^ Strict prevention and control measures can greatly reduce the scope of the epidemic; extensive surveillance and detection are extremely important in containing its spread.^28^ On this basis, we divided government governance and prevention into seven evaluation indicators: government effectiveness, e-government, statistical capacity, information technology, emergency preparedness, stringency (a measure of the number and rigor of the measures taken), and monitoring and testing.

#### 2.2.2 Health Financing

Health financing is one of the key ways to support the development of a health system. Improved funding can drive improvements in the provision of primary health care and enable the system to respond effectively to changing population health needs and to mitigate shocks from public health emergencies.^29^ Thus, the monetary input of a country’s or region’s health system is particularly important for the emergency response of its health system. On this basis, we divided health financing into three evaluation indicators: national health expenditures, population health expenditures, and insurance coverage.

#### 2.2.3 Health Service Provision

Health service provision has been a common concern in all regions.^30^ The physical accessibility of health facilities is one of the most important factors affecting the use of health services by patients during a public health emergency.^15^ Universal health care attempts to improve the quality of the health system through effective health coverage.^31^ However, health service provision is still influenced by a variety of factors such as ease of access to basic health services and the level of health care. On this basis, we divided health service provision into six evaluation indicators: the number of health institutions, the number of hospital beds, medical access, health coverage, health care, and health service supply pressure.

#### 2.2.4 Health Workforce

Health workers face a larger and more stressful workload than usual during an epidemic, so they need to be more resilient and adapt quickly to the changing situation.^32^ Health workers are a fundamental part of the health system; they perform duties that include carrying out medical research with the aim of improving disease prevention, diagnosis and treatment, clinical consultation, and provision of care to safeguard each patient. When faced with a public health crisis, health workers are often the first to respond. Adequate mobilization and coordination of health workers and medical supplies can significantly reduce mortality.^26^ On this basis, we divided health workers into five evaluation indicators: the numbers of surgeons, physicians, nurses, pharmacists, and biomedical technicians.

### 2.3. Calculation Methodology

We started with a three-scale method to determine the weight of the four first-level indicators; in this method, the weight has three values that define whether an indicator’s value is equal to, less than, or greater than the weight of another indicator. We then used the entropy-weight method to determine the weights of the second-level indicators, and finally used TOPSIS. Based on the results, we performed hierarchical clustering to group the 60 countries based on their health system resilience. In the final step, we analyzed the first-level indicators for each country to determine the factors that most strongly promoted or decreased resilience. The calculation process is described in sections 2.3.1 to 2.3.4. Figure 3 provides an overview of these steps.

**Figure 3.**
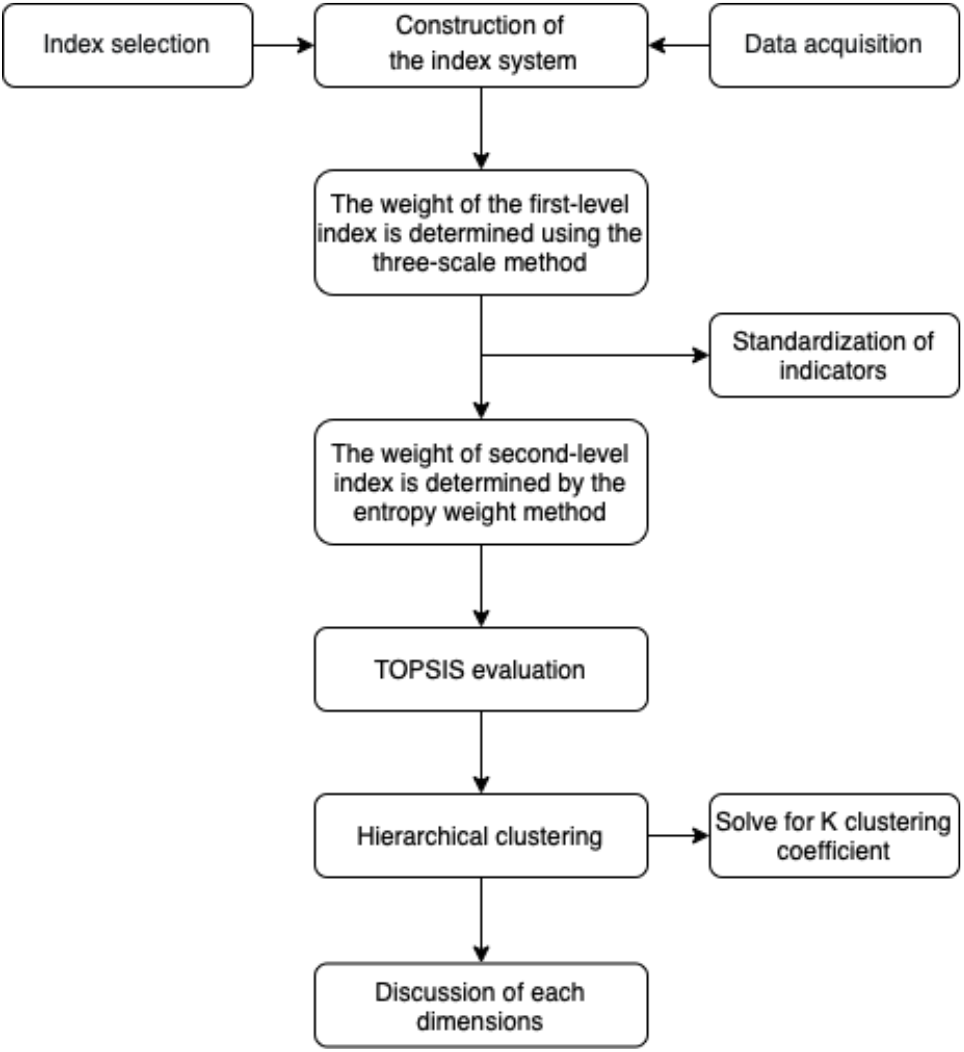
Methodology flow chart.

#### 2.3.1 Index Weighting

In this paper, we determined the final indicator weights by combining the three-scale method with the entropy-weighting method. This goes beyond following the subjective will of the decision makers to use objective goals and real data, thereby combining the benefits of subjectivity and objectivity. We used the three-scale method to determine the weights of the first-level indicators, followed by the entropy method to determine the weights of the second-level indicators under each first-level indicator. We obtained the final weights of the 21 secondary indicators by multiplying the first-level indicator weights by the proportional contribution of each second-level indicator to the total weight of the first-level indicator it supports.

The three-scale method is easy for experts to judge, makes it easy to construct a priority judgment matrix that summarizes these judgments, and provides a logical tool (the judgment matrix) for ranking priorities. Therefore, we used the three-scale method in this paper to determine the weight of the first-level indicators.^33^ (For more details, see Supplementary Text 2.1). Government governance and coordination are essential for any country, and at the heart of the global health and public service system is the prevention, detection, and response to public health threats.^34, 35^ When responding to an outbreak of infectious disease, timely government control and a tested emergency response plan can contain the outbreak most quickly. Therefore, government governance and prevention was the most important first-level indicator. Health workers are a fundamental part of the health system, but because they are trainable, responsive, and flexible, they are less important. Health financing is the driving force of the health system, but the input of health financing has a certain periodicity (e.g., because governments plan budgets annually), and it takes some time to see the effect of changes in budget, so we considered financing to be the second-most important first level indicator.^29^ Health service provision is an important component of the health system, since it requires the development of a health infrastructure, the provision of medical care, and the provision of basic health services. On this basis, we consider it the third-most important indicator.

The entropy weighting method constructs weights using the original (raw) data. The greater the variation in the data (i.e., differences between countries), the greater the amount of information contained in that indicator (i.e., the higher the entropy value) and the more important it is. We used the entropy method to objectively assess the weighting of the indicators. (Supplementary Section 2.2 describes details of the calculation for the second-level indicators.) We collected data for each indicator in the indicator system to construct the original (non-standardized) data matrix. The matrix contains data on the 21 second-level health system resilience indicators for 60 countries around the world. We standardized the indicators differently according to their characteristics. We then separately calculated the entropy value and weight of the second-level indicators. We then multiplied the weights of the first-level indicators by the proportional contribution of each second-level indicator to the corresponding first-level indicator, thereby obtaining the final weights of the 21 second-level indicators. (That is, the sum of these weights equaled the value of the corresponding first-level indicator.)

#### 2.3.2 Assessment of health system resilience by means of TOPSIS

We used the TOPSIS method to evaluate the decision matrix formed by each second-level indicator to obtain a ranking of the health system resilience of each country. TOPSIS evaluation is a multi-attribute decision-support method that is widely used in various areas of decision making or evaluation.^36^ In this paper, we use TOPSIS to evaluate the health system resilience matrix (**C**_*i*_) for the 60 countries and rank them from most to least resilient. (Supplemental Section S3 provides details.) The values of **C**_*i*_ are in the range (0,1); the higher the score (i.e., the closer to 1), the more resilient the country.

#### 2.3.3 Classification of health system resilience

We used version 26.0 of the SPSS software (https://www.ibm.com/analytics/spss-statistics-software) to perform hierarchical clustering of the health system resilience scores for the 60 countries. We used the contour coefficients to determine the optimal clustering scheme.

The first step is hierarchical clustering, which is a well-known clustering algorithm. Compared with *K*-means clustering, it offers the advantage of adaptive clustering, which does not require setting the number of classes (*K*) in advance, and the sample can be flexibly divided into different numbers of classes according to the actual needs of the analysis. Specifically, hierarchical clustering first regards each sample as a separate category and then calculates the minimum distance between pairwise samples. Next, the two classes with the smallest distance are merged into a single new class. Then the distance is recalculated between the new class and all other classes. This procedure iterates until all the samples finally merge into a single category.^15, 37^

The second step is to calculate the contour coefficient. The contour coefficient reflects the quality of the clustering scheme, and its value is in the range (–1, 1). The larger the contour value, the better the effect. The contour coefficient *S*(*i*) combines two factors, the degree of cohesion *a*(*i*) and the degree of dispersion *b*(*i*), to evaluate the clustering performance. For details of the calculation method, see Supplemental Text S4.

#### 2.3.4 Resilience of a Health Systems in Terms of the Four Dimensions

In this paper, we standardized the original (raw) data and multiplied the standardized values by the final weight of each second-level indicator that was obtained by the combined weighting method described in Supplemental Section S2. We then summed the values of each dimension to obtain the resilience score of each country under each dimension. Supplementary Text S5 provides details of the calculations. By comparing the health system resilience of 60 countries based on their responses to COVID-19 under the four first-level indicators, we obtained additional insights into the health system resilience of the 60 countries, which let us discuss the factors most strongly responsible for a country’s health system resilience and propose policy recommendations to improve its future performance.

## 3. Results

### 3.1 Ranking results

Table 1 presents the ranking of the health system resilience of the 60 countries. The 10 countries with the highest health system resilience were Switzerland, Japan, Germany, Australia, South Korea, Canada, New Zealand, Finland, the United States, and the United Kingdom. The 10 countries with the lowest health system resilience were (from best to worst) Liberia, Tanzania, Burundi, Mozambique, Nigeria, Benin, Côte d’Ivoire, Guinea, South Sudan, and Burkina Faso.

**Table 1.**
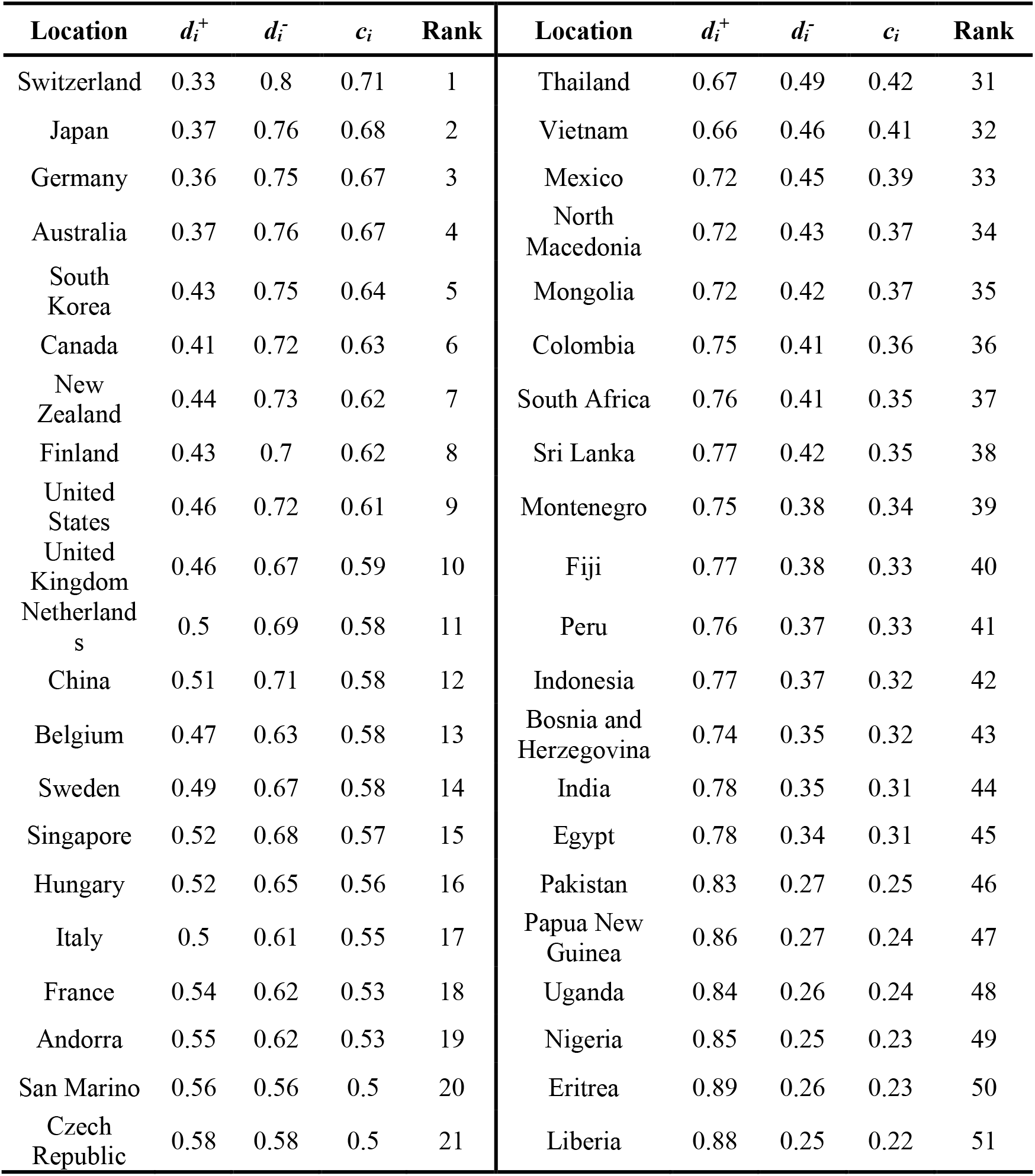
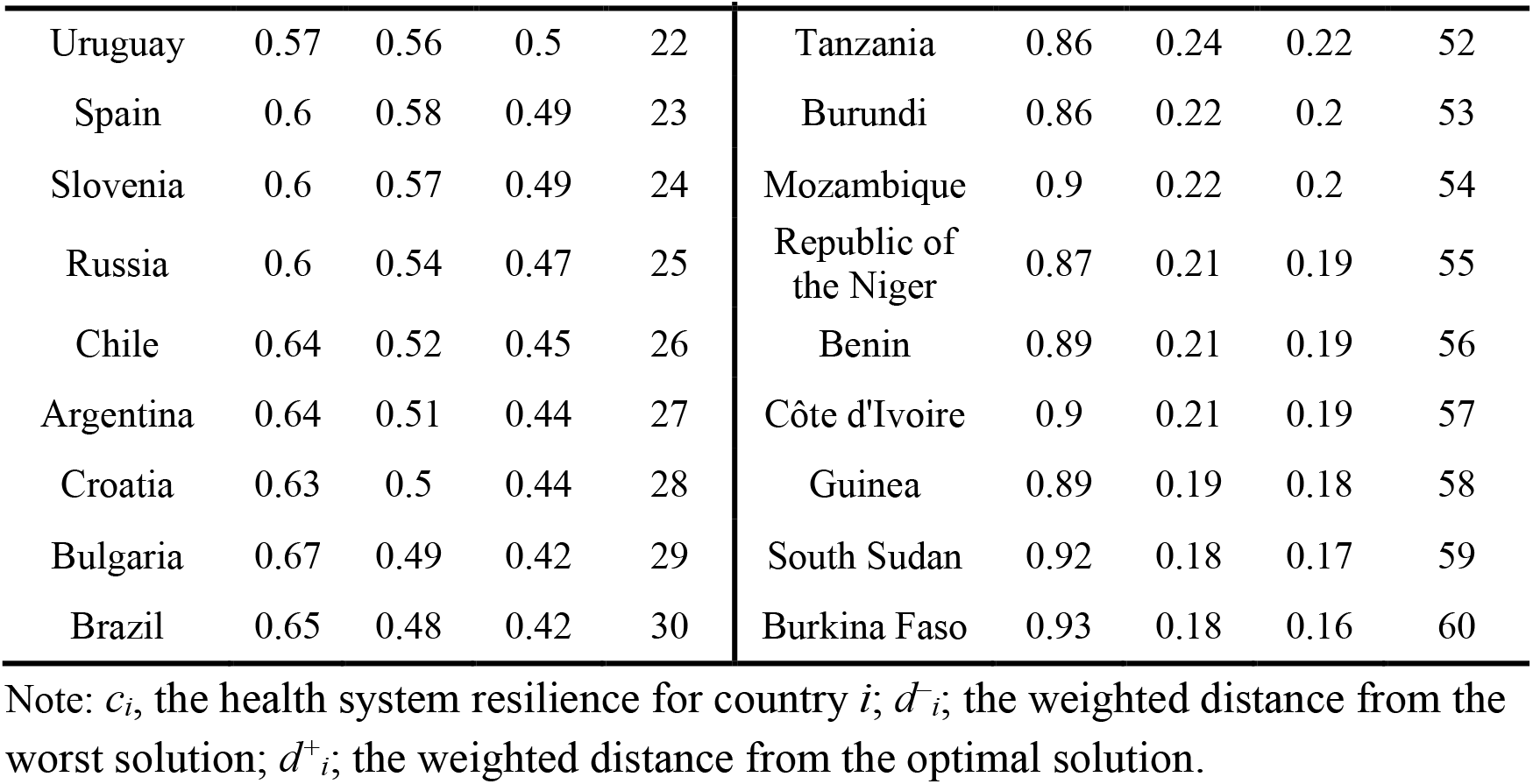
Global health system resilience rankings of the 60 countries.

| Location | $d_i^+$ | $d_i^-$ | $c_i$ | Rank | Location | $d_i^+$ | $d_i^-$ | $c_i$ | Rank |
| --- | --- | --- | --- | --- | --- | --- | --- | --- | --- |
| Switzerland | 0.33 | 0.8 | 0.71 | 1 | Thailand | 0.67 | 0.49 | 0.42 | 31 |
| Japan | 0.37 | 0.76 | 0.68 | 2 | Vietnam | 0.66 | 0.46 | 0.41 | 32 |
| Germany | 0.36 | 0.75 | 0.67 | 3 | Mexico | 0.72 | 0.45 | 0.39 | 33 |
| Australia | 0.37 | 0.76 | 0.67 | 4 | North Macedonia | 0.72 | 0.43 | 0.37 | 34 |
| South Korea | 0.43 | 0.75 | 0.64 | 5 | Mongolia | 0.72 | 0.42 | 0.37 | 35 |
| Canada | 0.41 | 0.72 | 0.63 | 6 | Colombia | 0.75 | 0.41 | 0.36 | 36 |
| New Zealand | 0.44 | 0.73 | 0.62 | 7 | South Africa | 0.76 | 0.41 | 0.35 | 37 |
| Finland | 0.43 | 0.7 | 0.62 | 8 | Sri Lanka | 0.77 | 0.42 | 0.35 | 38 |
| United States | 0.46 | 0.72 | 0.61 | 9 | Montenegro | 0.75 | 0.38 | 0.34 | 39 |
| United Kingdom | 0.46 | 0.67 | 0.59 | 10 | Fiji | 0.77 | 0.38 | 0.33 | 40 |
| Netherlands | 0.5 | 0.69 | 0.58 | 11 | Peru | 0.76 | 0.37 | 0.33 | 41 |
| China | 0.51 | 0.71 | 0.58 | 12 | Indonesia | 0.77 | 0.37 | 0.32 | 42 |
| Belgium | 0.47 | 0.63 | 0.58 | 13 | Bosnia and Herzegovina | 0.74 | 0.35 | 0.32 | 43 |
| Sweden | 0.49 | 0.67 | 0.58 | 14 | India | 0.78 | 0.35 | 0.31 | 44 |
| Singapore | 0.52 | 0.68 | 0.57 | 15 | Egypt | 0.78 | 0.34 | 0.31 | 45 |
| Hungary | 0.52 | 0.65 | 0.56 | 16 | Pakistan | 0.83 | 0.27 | 0.25 | 46 |
| Italy | 0.5 | 0.61 | 0.55 | 17 | Papua New Guinea | 0.86 | 0.27 | 0.24 | 47 |
| France | 0.54 | 0.62 | 0.53 | 18 | Uganda | 0.84 | 0.26 | 0.24 | 48 |
| Andorra | 0.55 | 0.62 | 0.53 | 19 | Nigeria | 0.85 | 0.25 | 0.23 | 49 |
| San Marino | 0.56 | 0.56 | 0.5 | 20 | Eritrea | 0.89 | 0.26 | 0.23 | 50 |
| Czech Republic | 0.58 | 0.58 | 0.5 | 21 | Liberia | 0.88 | 0.25 | 0.22 | 51 |
| Uruguay | 0.57 | 0.56 | 0.5 | 22 | Tanzania | 0.86 | 0.24 | 0.22 | 52 |
| Spain | 0.6 | 0.58 | 0.49 | 23 | Burundi | 0.86 | 0.22 | 0.2 | 53 |
| Slovenia | 0.6 | 0.57 | 0.49 | 24 | Mozambique | 0.9 | 0.22 | 0.2 | 54 |
| Russia | 0.6 | 0.54 | 0.47 | 25 | Republic of the Niger | 0.87 | 0.21 | 0.19 | 55 |
| Chile | 0.64 | 0.52 | 0.45 | 26 | Benin | 0.89 | 0.21 | 0.19 | 56 |
| Argentina | 0.64 | 0.51 | 0.44 | 27 | Côte d'Ivoire | 0.9 | 0.21 | 0.19 | 57 |
| Croatia | 0.63 | 0.5 | 0.44 | 28 | Guinea | 0.89 | 0.19 | 0.18 | 58 |
| Bulgaria | 0.67 | 0.49 | 0.42 | 29 | South Sudan | 0.92 | 0.18 | 0.17 | 59 |
| Brazil | 0.65 | 0.48 | 0.42 | 30 | Burkina Faso | 0.93 | 0.18 | 0.16 | 60 |

Note: $c_i$ , the health system resilience for country $i$ ; $d_i^-$ ; the weighted distance from the worst solution; $d_i^+$ ; the weighted distance from the optimal solution.

### 3.2 Clustering Results

Based on the contour coefficient calculations (Supplementary Figure S1), the optimal result was *K* = 3 clusters, which produced the highest contour coefficient (0.846). We therefore divided the 60 countries into three categories to discuss the resilience of their health systems. Figure 4 shows the cluster chart produced by the hierarchical cluster analysis.

**Figure 4.**
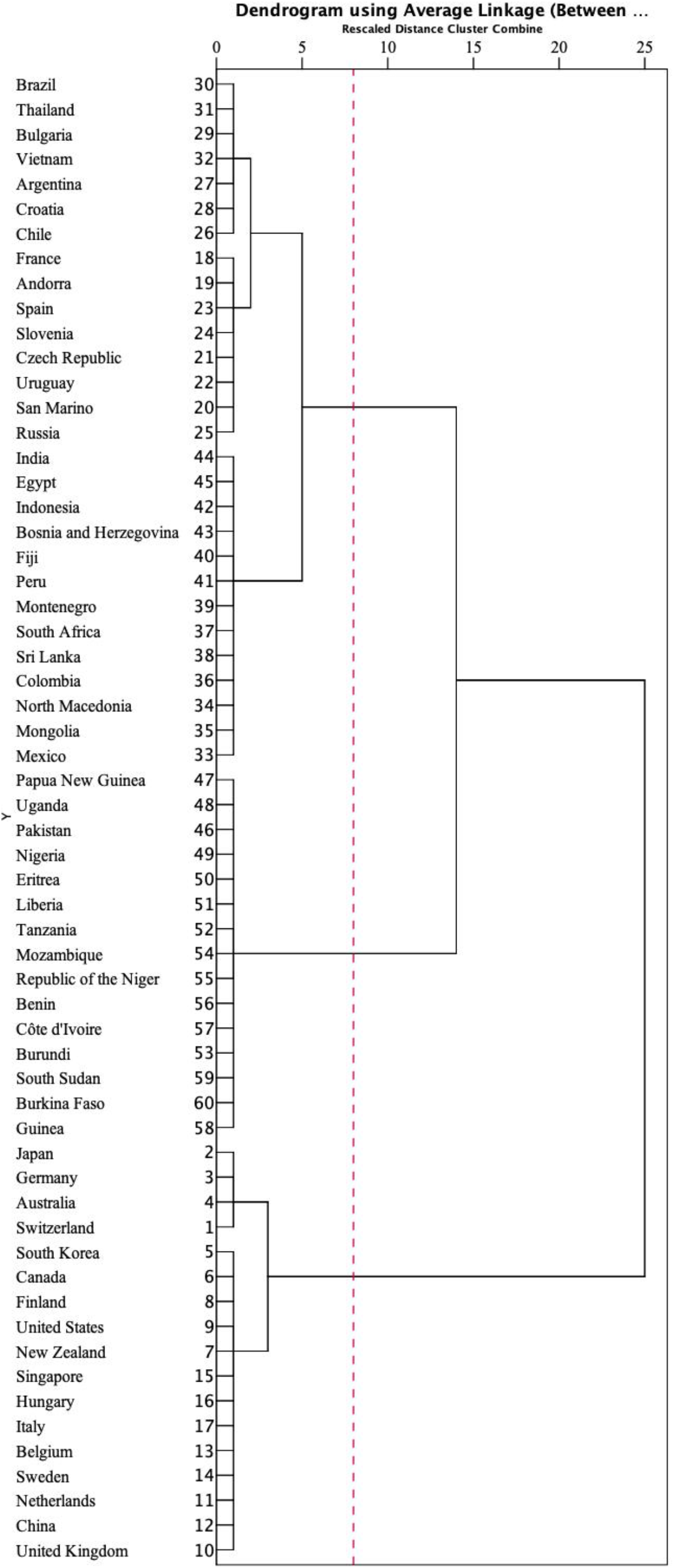
Clustering spectrum diagram.

Table 2 shows the division of the 60 countries into three categories: high, moderate, and low resilience.

**Table 2.**
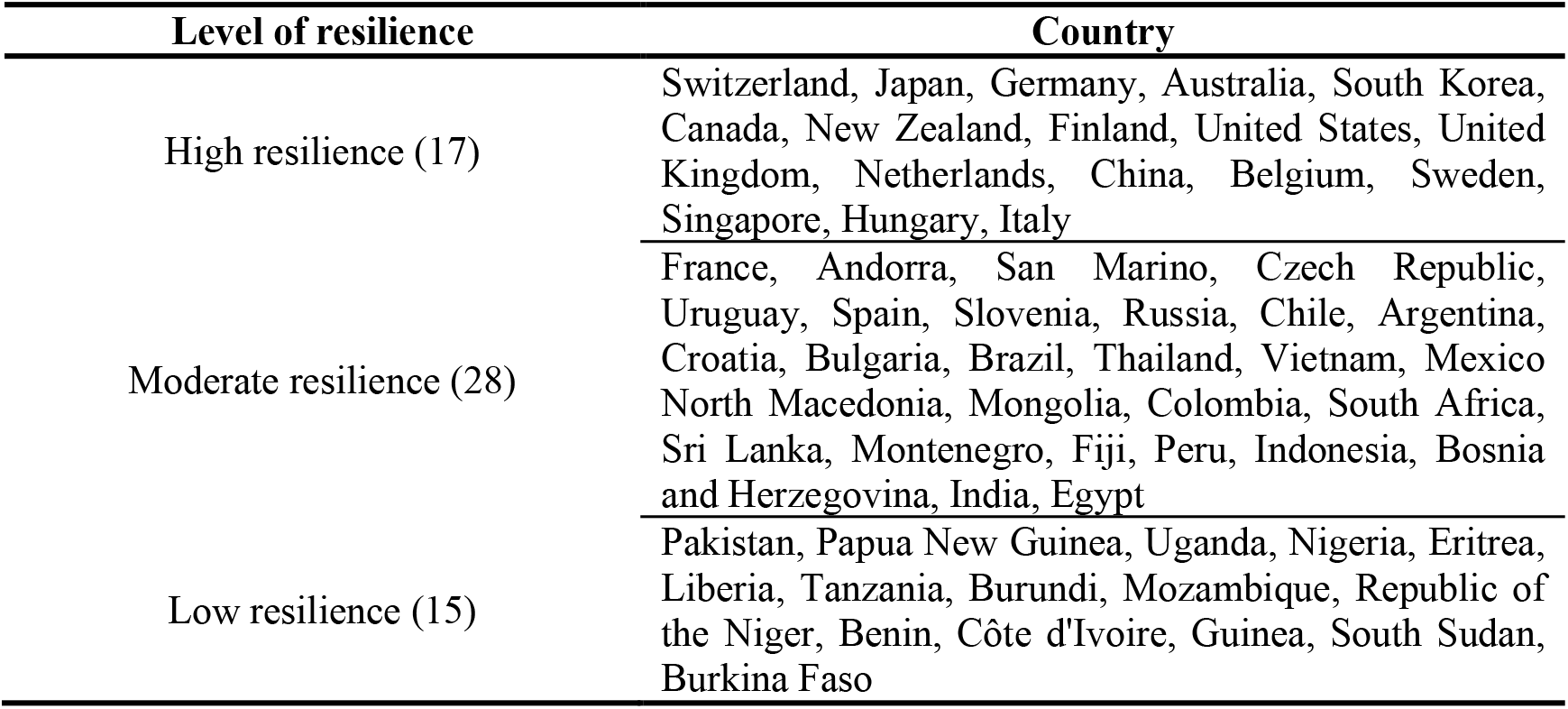
Ranking of health system resilience by the clustering analysis shown in Figure 4.

Next, we mapped the health system resilience values (Figure 5) to provide a visual representation of the global distribution of health system resilience. Most of the countries with high resilience are located in Western Europe, East Asia, North America. and Southern Oceania. Africa had a generally low resilience level, except for South Africa and Egypt, which had moderate resilience. Moderate resilience levels dominated South America, South Asia, and Eastern Europe.

**Figure 5.**
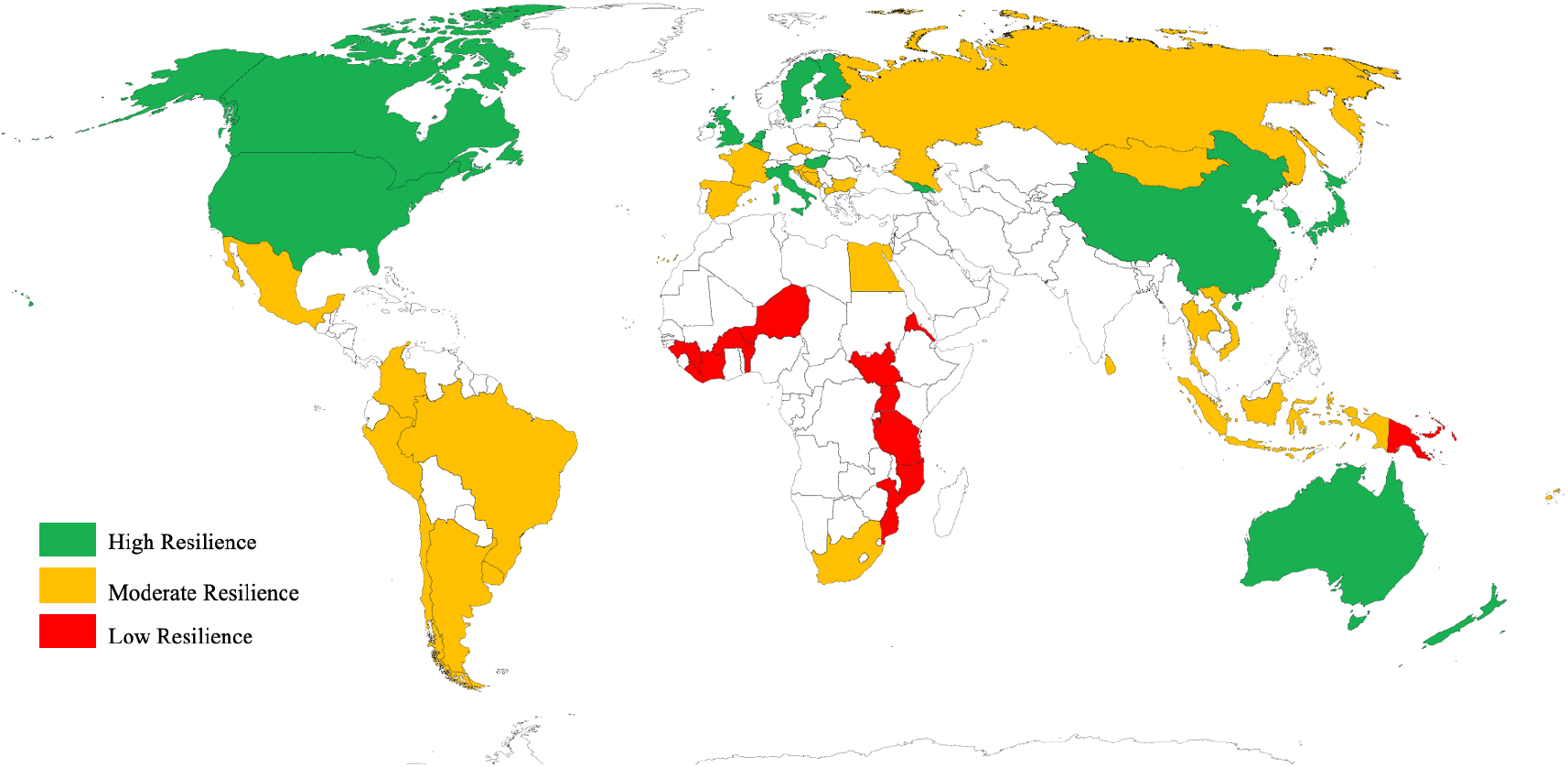
Global distribution of health system resilience.

### 3.3 Health System Resilience for the Four First-level Dimensions

Table 3 presents the scores for each of the four first-level indicators of resilience, with the cells colored to provide a visual summary of the resilience categories. The green color indicates a higher score and better resilience, yellow represents intermediate resilience, and red represents poor resilience. Note that each of the four dimensions has a different range of index values because of the different weights assigned to these dimensions. The government governance and prevention score averaged 0.257, versus 0.068 for health financing, 0.059 for health service provision, and 0.019 for health workers.

**Table 3.** Health system resilience scores in each dimension. Countries are presented in order of decreasing total score.

| Location | Government governance and prevention | Health Financing | Health service provision | Health workforce |
| --- | --- | --- | --- | --- |
| Switzerland[1] | 0.435 | 0.206 | 0.084 | 0.037 |
| Japan[2] | 0.433 | 0.124 | 0.113 | 0.053 |
| Germany[3] | 0.446 | 0.142 | 0.093 | 0.035 |
| Australia[4] | 0.455 | 0.136 | 0.100 | 0.028 |
| South Korea[5] | 0.453 | 0.090 | 0.122 | 0.021 |
| Canada[6] | 0.436 | 0.134 | 0.078 | 0.027 |
| New Zealand[7] | 0.445 | 0.116 | 0.080 | 0.026 |
| Finland[8] | 0.408 | 0.122 | 0.073 | 0.051 |
| United States[9] | 0.334 | 0.224 | 0.070 | 0.032 |
| United Kingdom[10] | 0.384 | 0.122 | 0.076 | 0.036 |
| Netherlands[11] | 0.386 | 0.136 | 0.071 | 0.023 |
| China[12] | 0.451 | 0.054 | 0.071 | 0.043 |
| Belgium[13] | 0.332 | 0.131 | 0.083 | 0.047 |
| Sweden[14] | 0.339 | 0.148 | 0.067 | 0.048 |
| Singapore[15] | 0.416 | 0.086 | 0.066 | 0.023 |
| Hungary[16] | 0.431 | 0.067 | 0.063 | 0.026 |
| Italy[17] | 0.356 | 0.100 | 0.072 | 0.045 |
| France[18] | 0.300 | 0.131 | 0.086 | 0.030 |
| Andorra[19] | 0.341 | 0.090 | 0.085 | 0.026 |
| San Marino[20] | 0.305 | 0.100 | 0.073 | 0.035 |
| Uruguay[21] | 0.316 | 0.080 | 0.078 | 0.017 |
| Czech Republic[22] | 0.309 | 0.080 | 0.080 | 0.033 |
| Spain[23] | 0.286 | 0.097 | 0.077 | 0.031 |
| Slovenia[24] | 0.303 | 0.087 | 0.070 | 0.031 |
| Russia[25] | 0.307 | 0.052 | 0.089 | 0.021 |
| Chile[26] | 0.276 | 0.077 | 0.058 | 0.027 |
| Argentina[27] | 0.258 | 0.075 | 0.068 | 0.021 |
| Croatia[28] | 0.257 | 0.066 | 0.069 | 0.029 |
| Thailand[29] | 0.275 | 0.047 | 0.070 | 0.010 |
| Brazil[30] | 0.255 | 0.072 | 0.055 | 0.023 |
| Bulgaria[31] | 0.247 | 0.058 | 0.065 | 0.030 |
| Vietnam[32] | 0.300 | 0.036 | 0.053 | 0.007 |
| Mexico[33] | 0.229 | 0.050 | 0.072 | 0.012 |
| North Macedonia[34] | 0.211 | 0.052 | 0.060 | 0.022 |
| Mongolia[35] | 0.205 | 0.039 | 0.072 | 0.023 |
| Colombia[36] | 0.188 | 0.057 | 0.055 | 0.011 |
| South Africa[37] | 0.193 | 0.064 | 0.047 | 0.006 |
| Sri Lanka[38] | 0.198 | 0.046 | 0.058 | 0.004 |
| Montenegro[39] | 0.187 | 0.032 | 0.061 | 0.007 |
| Fiji[40] | 0.184 | 0.046 | 0.044 | 0.005 |
| Peru[41] | 0.189 | 0.039 | 0.050 | 0.010 |
| Indonesia[42] | 0.219 | 0.026 | 0.035 | 0.003 |
| Bosnia and Herzegovina[43] | 0.172 | 0.050 | 0.051 | 0.013 |
| India[44] | 0.218 | 0.009 | 0.034 | 0.012 |
| Egypt[45] | 0.188 | 0.029 | 0.038 | 0.012 |
| Pakistan[46] | 0.164 | 0.013 | 0.029 | 0.004 |
| Papua New Guinea[47] | 0.087 | 0.030 | 0.039 | 0.001 |
| Uganda[48] | 0.167 | 0.013 | 0.022 | 0.001 |
| Nigeria[49] | 0.147 | 0.006 | 0.027 | 0.002 |
| Eritrea[50] | 0.103 | 0.017 | 0.024 | 0.001 |
| Liberia[51] | 0.046 | 0.038 | 0.036 | 0.010 |
| Tanzania[52] | 0.141 | 0.009 | 0.027 | 0.001 |
| Burundi[53] | 0.116 | 0.027 | 0.022 | 0.001 |
| Mozambique[54] | 0.086 | 0.022 | 0.022 | 0.001 |
| Republic of the Niger[55] | 0.116 | 0.015 | 0.022 | 0.000 |
| Benin[56] | 0.111 | 0.004 | 0.022 | 0.001 |
| Côte d'Ivoire[57] | 0.079 | 0.006 | 0.031 | 0.001 |
| Guinea[58] | 0.112 | 0.005 | 0.015 | 0.000 |
| South Sudan[59] | 0.039 | 0.023 | 0.025 | 0.002 |
| Burkina Faso[60] | 0.067 | 0.010 | 0.022 | 0.001 |

The 17 countries with a high resilience level had an average resilience score of 0.651. Most of these countries are developed countries with globally leading health systems, but there was significant variation in government governance, with values ranging from 0.332 to 0.455. The Swiss government responded strongly and effectively to the public health emergency by placing a high priority on surveillance and detection of COVID-19, with an indicator score of 0.435.^38^ Switzerland’s health financing indicator had an average score of 0.206, indicating a well-developed healthcare infrastructure and the most resilient health system based on its response to COVID-19.

Germany has one of the most advanced health care systems in the European countries, with a proactive health care strategy and the lowest mortality rate through early diagnosis and extensive detection, despite the fact that 21% of the population is over 65 years old (https://datacatalog.worldbank.org/dataset/world-development-indicators).

Australia, South Korea, Canada, and New Zealand, the four countries with the highest ranking for government governance, were able to contain the spread of the epidemic through effective outbreak prevention and control, and high levels of detection and tracking.^39^ However, health workers in these four countries scored low, with a mean score (0.025) that was only in the middle of the sample. This represented a high burden on health workers.

The US ranked first in health financing (0.224), with health expenditure accounting for 16.9% of domestic GDP and per capita health expenditure of US$10,623.9, with both values ranking first in the world. However, the US government ranked 17th in governance and prevention (0.334) and 26th in surveillance and detection, as the lack of a unified public health system led to poor coordination of surveillance and detection, making the epidemic worse.^40^ Similarly, the United Kingdom, which has a universal health care system and the highest health insurance coverage, ranked 13th in government governance and prevention (0.384), with low government emergency preparedness, lax prevention and control measures, and abrupt suspension of surveillance and detection, leaving the entire health system in distress.^41^

China and Hungary ranked high in Government Governance and Prevention, with both governments using their experience in efficient government governance and emergency preparedness to quickly contain the outbreak. However, the average health financing score in both countries was only 0.06, and there was a huge gap in health investment and a serious problem with uneven distribution of health resources.

Italy’s public health system has been adversely affected by privatization, with cuts of more than €37 billion in national healthcare services from 2010 to 2019, a continued decline in public health spending to 6.6% of GDP from 2018 to 2020, and an aging population, which was hit hard by COVID-19.^42^

There were 28 countries at the moderate resilience level, with an average resilience score of 0.388. These countries have a mid-range score for their health system, with some shortcomings and pre-existing problems within the country that were exacerbated by travel restrictions during the epidemic. France was 9th in health financing (with a score of 0.131) and has a high level of health investment, but its government stringency index was only medium, since the government took preventive and control measures only when the infection rate began to rise rapidly.^43, 44^

In terms of health service provision, Andorra (0.085), the Czech Republic (0.080) and Uruguay (0.078) have relatively robust health systems, with complementary public and private health care institutions, and a combination of clinics and mobile aid stations that provide multiple channels of care and protection for the population. Spain had one of the worst ratings for government governance and prevention in Europe, ranking 24th in emergency preparedness, with travel restrictions being implemented one week after the implementation of this policy in Italy.^45^ Surveillance and detection ranked 45th, with a very decentralized surveillance and detection system. Health financing (0.097) was below the average for developed countries, and decades of austerity measures have severely weakened the national health care system.^46^

Russian health financing (0.052) was below average (0.068). It ranked 46th on the stringency index, with lax epidemic control and lagging prevention and control measures that left the national health system in crisis. The rating of Russia’s health workforce (0.021) was only medium, with a shortage of health workers, stressful workloads, and a shortage of protective supplies. Russian health workers were therefore 16 times more likely to die from COVID-19 than health workers in other countries.^47^

Chile’s health service provision (0.058) was close to the average (0.059), with a low density of health care facilities and hospital beds that left many patients without timely care. Argentina and Mexico are deep in debt, and the epidemic prompted both countries to endure double pressure from a combination of their debt with other economic factors (greatly decreased trade and increased medical expenses). Argentina adopted strict preventive and control measures and ranked in the top 6 in terms of the stringency index. The trade blockade created by the epidemic hit the domestic economy hard, leading to frequent social crises that exacerbated Argentina’s deep debt. Mexico’s health service provision (0.072) was above average (0.059) and the country had a high health insurance coverage ratio, but was also affected by the country’s debt crisis, making health financing unsustainable and leaving the Mexican health system struggling.

Brazil saw a significant boost in health financing following the health system reform between 2000 and 2014, with total health expenditure rising from 7.0% of GDP to 8.3% and per capita health expenditure increasing from US$263 in 2000 to US$947 in 2014.^48^ However, their health system reform is incomplete, and the provision of basic health services is unevenly distributed. There are significant disparities across the country.

Thailand has received global recognition for its healthcare services, having been ranked 6^th^ in the world.^49^ The government health department used integrated technology to quickly carry out surveillance, detection, tracing, quarantine, and other prevention and control measures to achieve early control of the outbreak.^50^

The Vietnamese government showed excellent leadership in responding to COVID-19, ranking in the top quarter of countries in terms of its emergency preparedness performance and its surveillance and detection performance. Vietnam initiated the first public health emergency response at the beginning of the outbreak, sealing the border and taking the initiative to take early measures beyond WHO recommendations to minimize the outbreak.^51^

South Africa has one of the best health systems in Africa, but faces significant import pressure due to its close ties with Europe (leading to entry of infected individuals into the country and the spread of infection) and lax preventive and control measures (due to a failure to prevent imported infections), with government governance and prevention (0.193) well below average (0.257).

Sri Lanka has a weak health system, with a health workforce rating (0.04) equal to only about 20% of the average and below-average health financing, but the government was able to reduce the pressure on the health system by taking strict and effective control measures to limit the epidemic’s spread.^52^ In Indonesia, the government delayed the public health emergency response to avoid a large economic impact, and as a result, had the highest number of confirmed cases in March 2020 in Southeast Asia. This exacerbated a significant imbalance in access to health services.^53^ The Indian government governance and prevention rating (0.218) resulted from reduced air travel and increased road traffic control, but its surveillance and detection equipment were limited and their implementation was dubious because the quality of the government data has been questioned.^54^

There were 15 countries in the low-resilience category, with an average resilience score of 0.149. All of these countries had health system indicators well below those of countries with good and medium resilience. As a result, the health systems of these countries had serious potential for collapse at any time under the shock of an epidemic. Pakistan and Uganda, which had the same level of health financing (0.013) and similar government governance and prevention (0.164 and 0.167, respectively), had severely uneven distribution of health facility coverage and of health resources. The vast majority of the population lacks access to health care.

Libya’s and South Sudan’s public health efforts were more difficult to coordinate as they were often embroiled in war and conflict, greatly diminishing the public health infrastructure and other social benefits. Continued humanitarian crises will significantly reduce the ability of both countries to coordinate their response to COVID-19.^55^ Their government governance and prevention scores were only about 10% of Switzerland’s, with the worst emergency preparedness and lax prevention and control measures. This put the entire country’s society and economy at risk if an epidemic spreads.

Tanzania’s health financing score was only 0.009. Even though its government is implementing the Health Sector Strategic Plan IV 2015-2020 to improve the health system, implementation has faced huge obstacles to health financing reform due to low income and low demand, and the lack of health investment will make it difficult to improve the coverage of public health services.^56, 57^

Nigeria (0.006), Benin (0.004), and Côte d’Ivoire (0.006) scored the lowest in health financing, with structural imbalances and problems such as a lack of basic health coverage. Under the impact of COVID-19, their health systems were overburdened and health care was placed under serious pressure.^40^ Basic health services are severely lacking and vulnerable groups are not covered by health care. The Republic of the Niger and Guinea did well in emergency preparedness and in surveillance and detection, but health workers scored lowest because of very low health care coverage, severe health care shortages, and an overburdened system. Burkina Faso was rated 0.067 in government governance and prevention and 0.022 in health service provision. The number of health facilities per capita and medical beds per capita were among the lowest in the world, so the country’s health system faces a high risk of collapse during an epidemic.

## 4. Conclusions and policy implications

In this study, we constructed an indicator system for evaluating health system resilience based on a country’s responses to COVID-19. The indicator system is based on a health system framework that uses four first-level indicators (government governance and prevention, health financing, health service provision, and health workforce) and a total of 21 second-level indicators. We determined the weight of each indicator for 60 representative countries using a weighting method that combined the three-scale method with an entropy-weighting method. We then ranked health system resilience using the TOPSIS method, and classified the countries into groups using hierarchical clustering. Finally, we analyzed details of the health systems of countries with different levels of resilience to identify potential policy recommendations. We found large differences in the resilience of national health systems around the world, but the causes of these differences resulted from differences in the four first-level indicators.

The regions with high health system resilience scores were mainly located in Western Europe, East Asia, North America and Southern Oceania, whereas the regions with low scores were mainly located in Africa; Eastern Europe, South America, and parts of Southeast Asia had intermediate resilience. The differences were strongly related to differences in economic and social development, exacerbated by differences in national healthcare strategies. The degree of government governance and prevention greatly influenced the response to the epidemic, which is mainly influenced by government emergency preparedness, the rigor of their responses (i.e., the stringency index), and their testing capability. Due to the lack of vaccines and specific treatments during the early stages of the 2020 epidemic, maintaining social distance and wearing a mask were the main approaches to resisting the COVID-19 epidemic.^58, 59, 60^ On the other hand, detection and isolation remain the main ways to detect and treat infectious diseases. These measures can greatly reduce mortality and control the spread of an epidemic (https://www.who.int/dg/speeches/detail/who-director-general-s-opening-remarks-at-the-media-briefing-on-covid-1918-march-2020).

Health financing was a core indicator of resilience, as national health spending determines the base level of health systems in society as a whole. Cuts in health financing have direct and difficult to reverse impacts on health systems. Low investment in health systems has left public health systems fragmented in South American countries. According to the UN^61^, COVID-19 exacerbated severe socioeconomic inequalities in South America and will force more than 45 million people into poverty.

In terms of health service provision, hospitals and intensive care unit beds played a key role in clinical care during the epidemic, but the role of primary health care systems and facilities was less prominent.^62^ Aging was a major factor in the higher mortality rates in developed countries. Health workers generally responded well to COVID-19, despite working long hours under exhausting and demoralizing conditions, but the lack of preparedness, facilities, vaccines, protective gear, and critical medicines left health workers in most countries in a precarious environment, with serious implications for their health and for health system stability.

Resilient health systems can respond effectively to the epidemic, providing strong national protection and mitigating the negative impact of the epidemic. Our results demonstrated that all four first-level indicators contributed to resilience, and that success in achieving resilience required high scores for all indicators; no single indicator was sufficient to compensate for low scores in other indicators. To achieve resilience, we have the following additional policy recommendations:

First, countries with higher levels of resilience, excellent health systems, and adequate medical resources have significant international responsibilities during the global fight against a pandemic. They should, while doing their best to prevent and control the domestic epidemic, play a leadership role in organizing the world’s response. They must show innovative global health development assistance, possibly using means such as telemedicine to help developing and less developed countries, must increase medical research investment and output, and must base their decisions on science. They must also learn from less-developed countries, many of which had better ratings in some resilience indicators.

Second, for countries with a moderate level of resilience, governments should devote their resources to identifying and correcting deficits in their health system, which is particularly difficult given the need to balance containment of the epidemic with economic stability. They must take full advantage of medical policy suggestions revealed by the responses of the developed countries to bridge the gap between research and policy. The government should rapidly formulate scientific epidemic prevention policies based on the best practices that were revealed during the epidemic. They must adjust domestic public health funding to ensure that the health system covers their entire population, including both COVID-19 treatment and follow-up treatment once someone has recovered from the initial infection. In addition, they must enhance their capacity to absorb and adapt to shocks, while maintaining social stability. Some countries that are fortunate in having more than adequate resources should allocate excess resources to support the international community and help it fight the pandemic.

Third, for countries with low resilience, governments should take the strictest prevention and control measures possible and adopt stricter requirements for personal protection to prevent the spread of the epidemic while still ensuring people’s ability to earn their livelihood as much as possible. Given constraints on their resources, they must allocate resources in the most effective ways. They should work with domestic and foreign nongovernmental organizations to maximize their roles. Governments must obtain good statistics so they understand the current state of the epidemic and its trend, and must communicate an accurate description of their situation to international organizations such as WHO. They must carefully manage their budget to allocate funds to the most important and effective social assistance and economic support programs, and seek aid from the international community to avoid the collapse of health systems and increased poverty and social inequalities that follow public health emergencies.

Compared with existing models, our approach offers several advantages. First, it relies on open official data sources, with relatively reliable data, relatively easy access to the data, and low update costs. Second, the combination of the three-scale method and the entropy-weighting method allowed both subjective assessment from experts (the three-score method) and objective assessment using the entropy-weighting method. Third, the TOPSIS method provides an objective way to improve the accuracy of the assessment. Fourth, hierarchical clustering provides an objective and powerful way to classify countries into groups so that planners can provide different types of assistance to different groups of countries, thereby providing more targeted response strategies.

Our approach has several limitations. Due to the time delay between database updates, our research results were based only on data from 2020. Because global vaccine data are not yet synchronized, we could not include this important indicator in our method to provide a more comprehensive assessment. Nonetheless, though the indicators in our framework are by no means exhaustive, they provided a realistic overview of how health systems around the world responded to the epidemic.

The COVID-19 epidemic has taken a huge toll around the world, and has exposed problems in national health systems around the world. There is an urgent need to reimagine and repair the broken global health system.^63^ Through assessments of health system resilience, problems can be identified so that targeted measures can be taken to remedy them. In particular, more attention needs to be paid to the allocation of global health resources. Collecting more information will help determine the priority countries that need the most support and explore new methods of providing health aid. Governments should prioritize countries and regions with less resilient health systems and help them to build a more resilient health system that can better respond to future public health emergencies. Only by improving the resilience of global health systems can we hope to respond successfully to future epidemics, improve human health, and move towards a healthier future.

## Data Availability

All data produced are available online at

## Funding

This work was supported by grants from the National Social Science Foundation of China (Project No. 18AZD005; No. 21BGL217), the National Natural Science Foundation of China (Project No. 71874108), and the Shanghai Soft Science Foundation of China (22692107800).

## Notes

### Competing Interest Statement

The authors have declared no competing interest.

