## Supplementary material for "Evaluation of health system resilience in 60 countries based on their responses to COVID-19": Evaluation of health system resilience in 60 countries based on their responses to COVID-19_Supplemental.docx

### ABSTRACT

**Introduction:** In 2020, the COVID-19 epidemic swept the world, and many national health systems faced serious challenges. To improve future public health responses, it’s necessary to evaluate the performance of each country’s health system.

**Methods**: We developed a resilience evaluation system for national health systems based on their responses to COVID-19 using four resilience dimensions: government governance and prevention, health financing, health service provision, and health workers. We determined the weight of each index by combining the three-scale and entropy-weight methods. Then, based on data from 2020, we used the Technique for Order of Preference by Similarity to Ideal Solution (TOPSIS) method to rank the health system resilience of 60 countries, then used hierarchical clustering to classify countries into groups based on their resilience level. Finally, we analyzed the causes of differences among countries in their resilience based on the four resilience dimensions.

**Results:** Switzerland, Japan, Germany, Australia, South Korea, Canada, New Zealand, Finland, the United States, and the United Kingdom had the highest health system resilience in 2020. Eritrea, Nigeria, Libya, Tanzania, Burundi, Mozambique, Republic of the Niger, Benin, Côte d’Ivoire, and Guinea had the lowest resilience. Government governance and prevention of COVID-19 will greatly affect a country’s success in fighting future epidemics, which will depend on a government’s emergency preparedness, stringency (a measure of the number and rigor of the measures taken), and testing capability. Given the lack of vaccines or specific drug treatments during the early stages of the 2020 epidemic, social distancing and wearing masks were the main defenses against COVID-19. Cuts in health financing had direct and difficult to reverse effects on health systems. In terms of health service provision, the number of hospitals and intensive care unit beds played a key role in COVID-19 clinical care.

**Conclusion:** Resilient health systems were able to cope more effectively with the impact of COVID-19, provide stronger protection for citizens, and mitigate the impacts of COVID-19. Our evaluation based on data from 60 countries around the world showed that increasing health system resilience will improve responses to future public health emergencies.

### Supplementary materials

##### S1. Resilience framework for national health systems in response to COVID-19

**Supplementary Table S1** Indicator description and data sources

| Primary indicator: Government governance and prevention | | | |
| --- | --- | --- | --- |
| Secondary indicators | Description | Data sources | Links |
| Government effectiveness | Reflects the quality of government policy formulation and implementation, the quality of civil servants and their freedom from political pressure, and the quality of public service provision. | Worldwide Governance Indicators | www.govindicators.org |
| E-government | The application of modern information technology and network technology by government departments to provide public services to the community is a reflection of the state's ability to govern. | United Nations E-Government Survey 2020 | https://publicadministration.un.org/egovkb/en-us/Reports/UN-E-Government-Survey-2020 |
| Statistical capacity | Refers to a country's ability to produce high-quality statistics that meet the needs of the country and its citizens. | World Development Indicators | https://datatopics.worldbank.org/world-development-indicators/stories/statistical-performance-indicators.html |
| Information technology | Indicates the level of national information technology development and promotes a high level of sharing of information resources across society | Measuring the Information Society Report 2017 | https://www.itu.int/net4/ITU-D/idi/2017/index.html#idi2017rank-tab |
| Emergency preparedness | Refers to the overall level of preparedness and experience of society in relation to national emergencies | COVID-19 Regional Safety Assessment | https://public.flourish.studio/visualisation/3529461/?utm_source=showcase&utm_campaign=visualisation/3529461 |
| Stringency index | Refers to the effective measures taken by the government to contain the further spread of the epidemic situation and to safeguard the national interest and the lives of the people. | Oxford COVID-19 Government Response Tracker | https://www.bsg.ox.ac.uk/research/research-projects/oxford-covid-19-government-response-tracker |
| Monitoring and testing | Refers to the scope of monitoring and detection, the efficiency of monitoring and detection, the diversity of technologies, and the complexity of each government region. | COVID-19 Regional Safety Assessment | https://public.flourish.studio/visualisation/3529461/?utm_source=showcase&utm_campaign=visualisation/3529461 |

| Primary indicator: Health Financing | | | |
| --- | --- | --- | --- |
| Secondary indicators | Principles | Data sources | Links |
| National health expenditure | The amount of money consumed by a country (or region) for health care services in a given period of time | World Health Organization Global Health Expenditure database | apps.who.int/nha/database |
| Population health expenditure | Refers to the amount of money consumed by the population for healthcare products and services per year | World Health Organization Global Health Expenditure database | apps.who.int/nha/database |
| Insurance coverage | This refers to the number of people covered by health insurance in each country as a percentage of the total number of all nationals. | International Labour Organization | https://www.ilo.org |

| Primary indicator: Health service provision | | | |
| --- | --- | --- | --- |
| Secondary indicators | Principles | Data sources | Links |
| Health institutions | Refers to the number of public and private health institutions established by law to carry out activities of diagnosis and treatment of diseases. | Global Health Observatory | https://www.who.int/data/gho/data/indicators |
| Hospital beds | Refers to the number of beds in public and private health facilities where patients are admitted for inpatient treatment. | Global Health Observatory | https://www.who.int/data/gho/data/indicators |
| Medical access | By counting age-standardized and risk-standardized mortality rates for 32 causes of illness considered appropriate for healthcare, measured on a scale from 0 (worst) to 100 (best) mortality rates. | Global Burden of Disease Study 2015 | http://ghdx.healthdata.org/record/global-burden-disease-study-2015-gbd-2015-healthcare-access-and-quality-index-based-amenable |
| Health coverage | Refers to the coverage of essential services based on the capacity and access to services for reproductive, maternal, newborn and child health, communicable diseases, non-communicable diseases, and tracking interventions for the general and most-vulnerable populations. | Global Health Observatory | https://www.who.int/data/gho/data/indicators |
| Health care | An estimate of the overall quality of the healthcare system, healthcare professionals, equipment, staff, doctors, costs, etc. | NUMBEO | https://www.numbeo.com/health-care/rankings_by_country.jsp |
| Health service supply pressure | The structure of health care resources is influenced by the age structure of the population and is therefore portrayed using the degree of aging of the population. It refers to the dynamics of a corresponding increase in the proportion of older people in the population due to a decrease in the number of younger people and an increase in the number of older people. | World Bank | https://data.worldbank.org/indicator |

| Primary indicator: Health workforce | | | |
| --- | --- | --- | --- |
| Secondary indicators | Principles | Data sources | Links |
| Surgeons | Refers to professionals who are responsible for diagnosing diseases treated by surgery and providing surgical treatment to patients. | Lancet Commission on Global Surgery | lancetglobalsurgery.org |
| Physicians | Refers to professionals who diagnose internal diseases and provide non-surgical treatment to patients. | World Health Organization's Global Health Workforce Statistics | apps.who.int/globalatlas |
| Nurses | Refers to the performance of a full clinical operation or a single specialized nursing activity related to health care. | World Health Organization's Global Health Workforce Statistics | apps.who.int/globalatlas |
| Pharmacists | A professional who is responsible for providing drug knowledge and pharmacy services and monitoring for drug interactions in several drugs prescribed by a doctor. | World Health Organization's Global Health Workforce Statistics | apps.who.int/globalatlas |
| Biomedical technicians | Refers to trained and qualified biomedical technicians who design, assess, regulate, acquire, maintain, manage, and train in the safe use of healthcare technology in health systems around the world. | World Health Organization's Global Health Workforce Statistics | apps.who.int/globalatlas |

##### S2. Indicator weights

In this paper, we determined the final indicator weights by combining the three-scale method (Section S2.1) with the entropy-weighting method (Section S2.2). This combination of subjectivity and objectivity accounts for the subjective wishes of decision makers and the performance of objective and real data. We used the three-scale method to determine the weights of the first-level indicators, and then used the entropy method to determine the weights of the second-level indicators under each first-level indicator. We obtained the final weights of the 21 second-level indicators by multiplying the first-level indicator weights by the proportional contribution of each second-level indicator to the weight of the first-level indicator:

| $w_{j}=\overline{W_{i}}\times w_{j}^{i}$ | (S2-1) |
| --- | --- |

where $\overline{W_{i}}$ denotes the value in the three-scale method to obtain the first $i$ (*i* = 1, ..., 4) weights of the four first-level indicators, and $w_{j}^{i}$ denotes the entropy weight obtained by the entropy-weighting method for the first *i* (*i* = 1, 2, 3, 4) of the first-level indicators under the *j* (*j* = 1, ..., 21) second-level indicators.

##### S2.1 Three-scale method to determine the weights of the first-level indicators

The three-scale method makes it easy for experts to make logical judgments about the relative importance of the indicator dimensions, so we adopted the three-scale method to determine the weights of the first-level indicators. The specific steps in the three-scale method are as follows:

The first step is to construct the characteristic factor judgment scale matrix. In the three-scale method, the set of values for each comparison of indicators is (0, 1, 2), and the comparison is done for each pair of indicators to build a comparison matrix. In that matrix, each indicator **D** = (*d_ij_*) = [0, 1, 2] denotes the set of judgement scales, and *d_ij_* = 0 means that indicator *i* is less important than indicator *j*, *d_ij_* = 1 means that the two indicators are equally important, and *d_ij_* = 2 means that indicator *i* is more important than indicator *j*. The results of this comparison are shown in supplementary Table S2.

Supplementary Table S2 Relative values of the four first-level indicators of resilience.

| Indicator (*i*) | *Ggp* | *Hf* | *Hsp* | *Hw* | *b_i_* |
| --- | --- | --- | --- | --- | --- |
| *Ggp* | 1 | 2 | 2 | 2 | 7 |
| *Hf* | 0 | 1 | 2 | 2 | 5 |
| *Hsp* | 0 | 0 | 1 | 2 | 3 |
| *Hw* | 0 | 0 | 0 | 1 | 1 |

Note: *Ggp is* government governance and prevention and control, *Hf is* health financing, *Hsp is* health service provision, *Hw* is health workers, and *b_i_* is the sum of each row of the matrix.

The second step is to determine the eigenfactor judgment matrix. We constructed the judgment matrix using the polar difference method. If *y_­ij_* denotes eigenfactor *i* and the ratio of the importance of eigenfactor *j* the ratio of the importance of the eigenfactors, then the eigenfactor *j* is the ratio of the importance of *i* to the ratio of the importance of 1/*y_ij_* and the extreme difference method f(*b_i_*, *b_j_*) $=y_{ij}=y_{b}^{\frac{b_{i}-b_{j}}{B}}.$ The resulting matrix **Y** = (*y_ij_*) is the consistency judgment matrix, and *y*_b_ is the relative importance of the pairs of polar-difference elements pre-given according to some criterion, and is a constant, generally taken as *y*_b_ = 9 (Liu et al. 2015). *B* = max(*b*_1_, *b*_2_, *b*_3_, *b*_4_) – min(*b*_1_, *b*_2_, *b*_3_, *b*_4_) is the polar difference. In this paper, we chose *B* = 6 as the extreme difference. Accordingly, the judgment matrix of characteristic factors is obtained:

| $\mathbf{z}=\left( y_{ij} \right)=\left[ \begin{matrix} y & \mathrm{Ggp} & \mathrm{Hf} & \mathrm{Hsp} & \mathrm{Hw} & M_{i} & W_{i} & \bar{W_{i}} \\ \mathrm{Ggp} & 1 & 2.080 & 4.327 & 9 & 81 & 3 & 0.542 \\ \mathrm{Hf} & 0.481 & 1 & 2.080 & 4.327 & 4.327 & 1.277 & 0.230 \\ \mathrm{Hsp} & 0.231 & 0.481 & 1 & 2.080 & 0.231 & 0.783 & 0.141 \\ \mathrm{Hw} & 0.111 & 0.231 & 0.481 & 1 & 0.012 & 0.481 & 0.087 \end{matrix} \right]$ | (S2-2) |
| --- | --- |

Where *Ggp is* government governance and prevention and control, *Hf is* health financing, *Hsd is* health service provision, *Hw* is health workers, and the top row of the table represents indicator *i* and the first column represents indicator *j*. In addition:

| $M_{i}=\prod_{j=1}^{4} y_{ij}$ | (S2-3) |
| --- | --- |
| $W_{i}=\sqrt[4]{M_{i}}$ | (S2-4) |
| $\sum_{i=1}^{4} W_{i}=5.541$ | (S2-5) |
| $W_{i}=\frac{W_{i}}{\sum_{j=1}^{4} W_{j}}(i=1, 2, 3, 4)$ | (S2-6) |
| $\sum_{i=1}^{4} W_{i}=1$ | (S2-7) |

The transpose matrix of the four first-level indicators, **W**^T^ = {0.542, 0.230, 0.141, 0.087} is the weight of the four first-level indicators.

The third step is a consistency test. Let **C** be the first four columns of the eigenfactor judgment matrix, and let **L** = (*l_i_*)_4×1_ = **C**×**W**^T^ = (2.413, 1.160, 0.558, 0.268). The largest characteristic root λ_max_ $=\frac{1}{4}\sum_{i=1}^{4} \frac{l_{i}}{W_{i}}=4$ , *P*_CI_ = (λ_max_ – 4)/(4–1) = 0 ≤ ε (ε = 0.001), where *P*_CI_ represents the index of consistency and ε represents the level of significance, and satisfies the consistency test.

The weights for the four first-level indicators are therefore:

**W**^T^ = (*W*_1,_ *W*_2,_ *W*_3,_ *W*_4_) = (0.542, 0.230, 0.141, 0.087)

##### S2.2 Entropy-weighting method to determine the weights of the second-level indicators

The entropy-weighting method constructs indicator weights using the original (non-standardized) data. Thus, we used the entropy weighting method to objectively assess the indicator weights. The entropy weighting method determines the weights of the second-level indicators as follows:

First, we constructed a decision matrix for evaluation of the health system resilience. We collected data for each indicator in the indicator system and constructed the original data matrix A, which contains data for *m* = 60 countries under *n* = 21 second-level health system resilience evaluation indicators.

| $\mathbf{A}=\left\{ \begin{matrix} a_{11} & \cdots& a_{1j} & \cdots& a_{1,21} \\ \vdots& \vdots& \vdots& \vdots& \vdots\\ a_{i1} & \cdots& a_{ij} & \cdots& a_{i,21} \\ \vdots& \vdots& \vdots& \vdots& \vdots\\ a_{60,1} & \cdots& a_{60,j} & \cdots& a_{60,21} \end{matrix} \right\}=\left( a_{ij} \right)_{60\times21}$ | (S2-8) |
| --- | --- |

The health system resilience indicators (*a_ij_*) are treated differently based on the nature of the indicators:

For revenue-based indicators:

| $x_{ij}=\left\{ \begin{aligned} \frac{a_{ij}-m_{j}^{-}}{m_{j}^{+}-m_{j}^{-}}, \#m_{j}^{+}\neq m_{j}^{-}\#\# \\ 1, \#m_{j}^{+}=m_{j}^{-} \end{aligned} \right.$ | (S2-9) |
| --- | --- |

For cost-based indicators:

| $x_{ij}=\left\{ \begin{aligned} \frac{m_{j}^{+}-a_{ij}}{m_{q}^{+}-m_{q}^{-}},\#m_{q}^{+}\neq m_{q}^{-} \\ 1,\#m_{q}^{+}=m_{q}^{-} \end{aligned} \right.$ | (S2-10) |
| --- | --- |

where $m_{j}^{+}$ denotes the maximum value of column *j*; that is, $m_{j}^{+}=\max\left( a_{ij} \right), i=1,2,\ldots,60. m_{j}^{-}$ denotes the minimum value of column *j*; that is*,* $m_{j}^{-}=\min\left( a_{ij} \right), i=1,2,\ldots,60$.

The decision matrix X is then obtained as follows:

| $\mathbf{X}=\left\{ \begin{matrix} x_{11} & \cdots& x_{1j} & \cdots& x_{1,21} \\ \vdots& \vdots& \vdots& \vdots& \vdots\\ x_{i1} & \cdots& x_{ij} & \cdots& x_{i,21} \\ \vdots& \vdots& \vdots& \vdots& \vdots\\ x_{60,1} & \cdots& x_{60,j} & \cdots& x_{60,21} \end{matrix} \right\}=\left( x_{ij} \right)_{60\times21}$ | (S2-11) |
| --- | --- |

Next, decision matrix X is standardized and transformed into dimensionless data to give the standardized matrix X’:

| $\mathbf{X}^{'}=\left\{ \begin{matrix} x_{11}^{'} & \cdots& x_{1j}^{'} & \cdots& x_{1,21}^{'} \\ \vdots& \vdots& \vdots& \vdots& \vdots\\ x_{i1}^{'} & \cdots& x_{ij}^{'} & \cdots& x_{i,21}^{'} \\ \vdots& \vdots& \vdots& \vdots& \vdots\\ x_{60,1}^{'} & \cdots& x_{60,j}^{'} & \cdots& x_{60,21}^{'} \end{matrix} \right\}=\left( x_{ij}^{'} \right)_{60\times21}$ | (S2-12) |
| --- | --- |

Where:

| $x_{ij}^{'}=\frac{x_{ij}}{\sum_{t=1}^{60} x_{tj}},i=1,2,\cdots,60;j=1,2,\cdots,21$ | (S2-13) |
| --- | --- |

Next, the entropy values of the second-level indicators (*e_j_*) under each first-level indicator and their weights (*w_j_*) are calculated separately:

| $e_{j}=-\frac{1}{\ln60}\sum_{i=1}^{60} \left( x_{ij}^{'}\times\ln x_{ij}^{'} \right),\left( j=1,\cdots,21 \right)$ | (S2-14) |
| --- | --- |

| $w_{j}^{i}=\frac{1-e_{j}}{\sum_{I_{j}\in B_{i}} \left( 1-e_{j} \right)}，\left( j=1,\ldots,21 \right)$ | (S2-15) |
| --- | --- |
| $\sum_{I_{j}\in B_{i}} w_{j}^{i}=1，\left( j=1,\ldots,21 \right)$ | (S2-16) |

The set of indicators corresponding to the data matrix is {*I*_1_, *I*_2_, …, *I_j_*, … *I*_21_} and **B***_i_* is the set of second-level indicators corresponding to *i*. If the indicators in the column of the data matrix (i.e., the second-level indicators) belong to the set, then: -th first-level indicator $I_{j}$. If the indicators in the column of the data matrix (second-level
indicators) belong to the set, then:

Where $w_{j}^{i}$ denotes the weight of the second-level indicators corresponding to first-level indicator *i*, and the indicators in column *I_j_* (second-level indicators) belong to set **B***_i_*.$\sum_{I_{j}\in B_{i}} w_{j}^{i}$ represents the sum of the weights of all second-level indicators under first-level indicator *i*.

In summary, the final weights of the 21 secondary indicators (*w_j_*) can be obtained using equation S2-1, which was defined at the start of Section S2.

##### S3. Assessment of health system resilience using TOPSIS

We used the TOPSIS method to evaluate the decision matrix formed by each second-level indicator and obtain a ranking of the health system resilience of each country. TOPSIS used the following calculation process:

Using the standardized decision matrix X’ = (*x*’*_ij_*)_60×21_ and the indicator weights, we calculated the weighted values of each indicator (Z):

| $\mathbf{Z}=\left( z_{ij} \right)_{60\times21}=\left\{ \begin{matrix} {w_{1}x}_{11}^{'} & \cdots& w_{21}x_{1,21}^{'} \\ \vdots& \ddots& \vdots\\ {w_{1}x}_{60,1}^{'} & \cdots& w_{21}x_{60,21}^{'} \end{matrix} \right\}$ | (S3-1) |
| --- | --- |

where *z_ij_* = *x*’*_ij_* × *w_j_*.

First, we find the largest number in each column, which is the ideal optimal solution vector (**Z**^+^):

| $\mathbf{Z}^{+}=\left( z_{1}^{+},\cdots,z_{21}^{+} \right),z_{j}^{+}=\max_{1\leq i\leq60}\left\{ z_{ij} \right\} (j=1, 2,\ldots, 21)$ | (S3-2) |
| --- | --- |

Next we find the smallest number in each column, which is the ideal worst solution vector (**Z**^–^):

| $\mathbf{Z}^{-}=\left( z_{1}^{-},\cdots,z_{21}^{-} \right),z_{j}^{-}=\min_{1\leq i\leq60}\left\{ z_{ij} \right\} (j=1, 2,\ldots, 21)$ | (S3-3) |
| --- | --- |

Next, for the first $i$ evaluation objects, we calculate the weighted distance from the optimal solution (*d^+^_i_*):

| $d_{i}^{+}=\sqrt{\sum_{j=1}^{21} \left( z_{j}^{+}-z_{ij} \right)^{2}}$ | (S3-4) |
| --- | --- |

and its weighted distance from the worst solution (*d^–^_i_*):

| $d_{i}^{-}=\sqrt{\sum_{j=1}^{21} \left( z_{j}^{-}-z_{ij} \right)^{2}}$ | (S3-5) |
| --- | --- |

Finally, we define the resilience index for country *i* as *c_i_*:

| $c_{i}=\frac{d_{i}^{-}}{d_{i}^{+}-d_{i}^{-}}(i=1, 2,\cdots, m)$ | (S3-6) |
| --- | --- |

Using this equation, we calculated the value of health system resilience for each of the 60 countries (*c_i_*) and ranked them. *c_i_* takes values in the range (0, 1). The smaller the value of *c_i_*, the smaller the distance between this evaluation object and the optimal solution; correspondingly, the smaller the $d_{i}^{-}$, the smaller the distance between this evaluation object and the worst solution. The evaluation result takes into account the distance between the optimal solution and the worst solution. The higher the score (i.e., the closer it is to 1), the closer the object is to the optimal level, and the more resilient the health system is.

##### S4. Classification of health system resilience by hierarchical clustering

We used version 26.0 of the SPSS software (https://www.ibm.com/analytics/spss-statistics-software) to perform hierarchical clustering of the health system resilience scores for the 60 countries. We used the contour coefficients to identify the best clustering scheme and classify the health system resilience levels of countries around the world and facilitate analysis of the response to different resilience levels. The specific steps are as follows:

The first step is hierarchical clustering, which starts by treating each sample as a separate class and calculating the minimum distance between each pair of samples. The software then merges the two classes with the smallest distance into a new class, and recalculates the distance between the new class and all remaining classes. The process iterates until all samples are finally combined into a single class.

In the second step, we calculate the contour coefficient, which reflects the quality of the clustering scheme and has a value within the range (–1, 1), with higher values indicating better results. The contour coefficient *S*(*i*) combines two factors to assess clustering performance: the cohesiveness *a*(*i*) and the dispersion *b*(*i*). For object *i*, cohesiveness indicates the average distance between other objects in the same cluster, whereas dispersion represents the average distance between object $i$ and all other objects in the cluster, where "distance" represents the degree of dissimilarity. The larger the contour coefficient, the more compact the objects are within a cluster and the greater the inter-cluster distance.

| $S\left( i \right)=\frac{\left[ b\left( i \right)-a\left( i \right) \right]}{\max\{a\left( i \right),b\left( i \right)\}}$ | (S4-1) |
| --- | --- |

Figure S1 shows the results of the clustering analysis. The highest contour coefficient was 0.846, and was produced with *k* = 3 clusters (i.e., the optimal number of classes was 3).

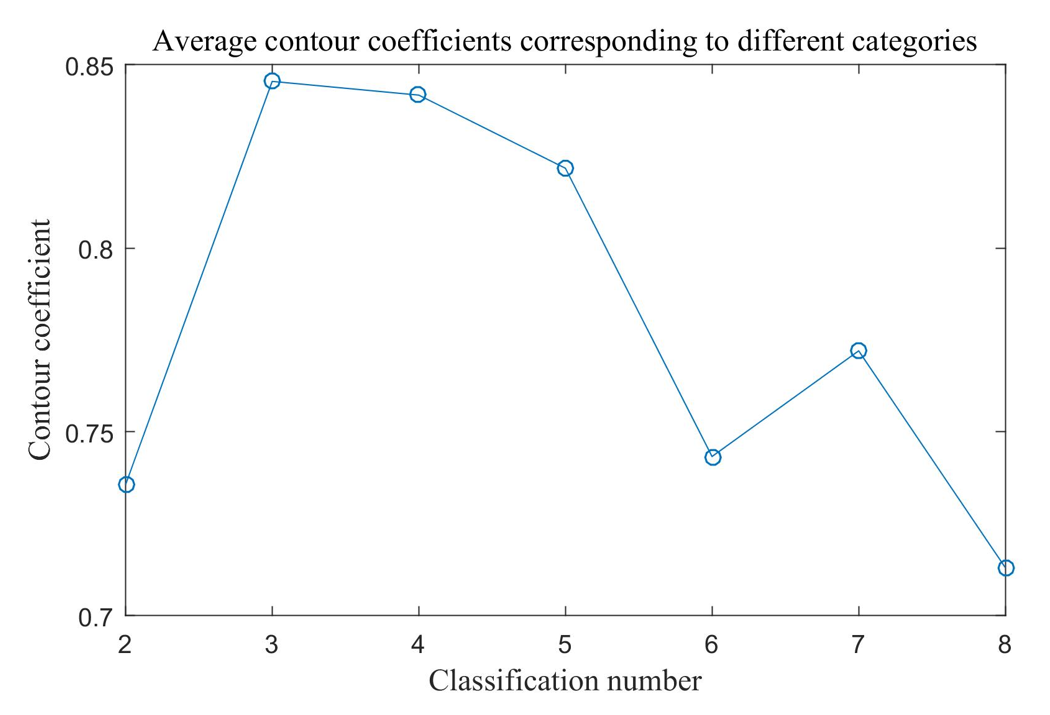

**Supplementary Figure S1.** Contour coefficients (*S*[*i*]) for different numbers of clusters (*K*).

We therefore divided the 60 countries into three categories to discuss the resilience of their health systems.

##### S5. Analysis under different dimensions

To further compare the magnitude of the health system resilience in the 60 countries, we examined the four first-level indicators. First, we standardized the original (non-standardized) data using equations S5-1 and S5-2, multiplied by the final weights (*w_j_*) of the second-level indicators calculated from equation 3-15$.$ The values for each dimension are then summed to obtain a resilience score for each country under each dimension. We then plotted the resilience scores for each dimension in a color-coded table to provide a more intuitive quantitative analysis. We then discussed the causes of the uneven development of health systems in each country.

For revenue-based indicators.

| $x_{ij}=\left\{ \begin{aligned} \frac{a_{ij}-m_{j}^{-}}{m_{j}^{+}-m_{j}^{-}}, \#m_{j}^{+}\neq m_{j}^{-}\#\# \\ 1, \#m_{j}^{+}=m_{j}^{-} \end{aligned} \right.$ | (S5-1) |
| --- | --- |

For cost-based indicators.

| $x_{ij}=\left\{ \begin{aligned} \frac{m_{j}^{+}-a_{ij}}{m_{q}^{+}-m_{q}^{-}},\#m_{q}^{+}\neq m_{q}^{-} \\ 1,\#m_{q}^{+}=m_{q}^{-} \end{aligned} \right.$ | (S5-2) |
| --- | --- |
